# High-throughput fetal-fraction amplification increases analytical performance of noninvasive prenatal screening

**DOI:** 10.1101/2020.07.12.20034926

**Authors:** Noah C. Welker, Albert K. Lee, Rachel A.S. Kjolby, Helen Y. Wan, Mark R. Theilmann, Diana Jeon, James D. Goldberg, Kevin R. Haas, Dale Muzzey, Clement S. Chu

**Affiliations:** Myriad Genetics, Inc. Salt Lake City, UT; Myriad Women’s Health, South San Francisco, CA

**Keywords:** fetal fraction, cell-free DNA, noninvasive prenatal screening, analytical validation, body mass index

## Abstract

**Purpose:** The percentage of a maternal cell-free DNA (cfDNA) sample that is fetal-derived (the fetal fraction; FF) is a key driver of the sensitivity and specificity of noninvasive prenatal screening (NIPS). On certain NIPS platforms, >20% of women with high body-mass index (and >5% overall) receive a test failure due to low FF (<4%).

**Methods:** A scalable fetal-fraction amplification (FFA) technology was analytically validated on 1,264 samples undergoing whole-genome sequencing (WGS)-based NIPS. All samples were tested with and without FFA.

**Results:** Zero samples had FF<4% when screened with FFA, whereas 1 in 25 of these same patients had FF<4% without FFA. The average increase in FF was 3.9-fold for samples with low FF (2.3-fold overall) and 99.8% had higher FF with FFA. For all abnormalities screened on NIPS, z-scores increased 2.2-fold on average in positive samples and remained unchanged in negative samples, powering an increase in NIPS sensitivity and specificity.

**Conclusions:** FFA transforms low-FF samples into high-FF samples. By combining FFA with WGS-based NIPS, a single round of NIPS can provide nearly all women with confident results about the broad range of potential fetal chromosomal abnormalities across the genome.

## INTRODUCTION

Since its introduction into clinical care nearly a decade ago,^1-5^ noninvasive prenatal screening (NIPS) based on cell-free DNA (cfDNA) has provided millions of pregnant women with information about their risk for fetal chromosomal abnormalities. A primary driver of NIPS sensitivity for aneuploidy in a given maternal plasma sample is the fetal fraction (FF), which describes the proportion of cfDNA fragments that originate from the placenta.^6^ For most samples, FF values are between 4% and 30%.^7^ Many laboratories fail samples with FF<4% to diminish the risk of issuing false-negative reports. Because the molecular and bioinformatic implementations of NIPS have evolved, diversified, and generally improved over time, sensitivity at progressively lower FF levels is platform- and laboratory-dependent.^6,8^ Indeed, a recently published clinical-experience study demonstrated that a customized whole-genome-sequencing (WGS)-based NIPS, which does not fail low-FF samples, can have comparable accuracy at high-FF and low-FF for the common aneuploidies on chromosomes 13, 18, and 21.^9^

Though the common aneuploidies have long been the main focus of NIPS because of their frequency and highly penetrant phenotype, clinically actionable chromosomal anomalies span a range of sizes and can occur anywhere in the genome.^10-12^ As such, a key frontier in NIPS development is to increase the resolution (i.e., detect smaller anomalies) and the scope (i.e., the number of regions) of the screen.

An example of increased resolution in NIPS is the screening for pathogenic microdeletion syndromes.^10,13,14^ such as DiGeorge Syndrome ^15^ and Cri du Chat Syndrome;^16^ these arise from deletions of megabases of genome sequence, which are detectable to varying degrees on the primary NIPS platforms (i.e., WGS-based,^17,18^ single-nucleotide-polymorphism (SNP)-based,^19,20^ and microarray-based^21^ platforms).

An example of increased scope in NIPS is screening for whole-chromosome aneuploidies on chromosomes other than 13, 18, and 21, also referred to as rare autosomal aneuploidies (RAAs), which are associated with pregnancy complications^12,22^ and can now be discovered with WGS-based NIPS.^12,17^ Each NIPS platform has the potential to achieve higher resolution (e.g., via more WGS depth, more probed SNPs, or more microarray probes); however, because only the WGS-based approach to NIPS intrinsically interrogates the whole genome, it is manifestly better suited to increase the scope of screening than targeted approaches like array- and SNP-based NIPS.

WGS-based NIPS has recently been configured to identify novel microdeletions anywhere in the genome, though in the one peer-reviewed characterization of such an offering, novel deletions must exceed 7MB in length.^17^ Since many pathogenic microdeletions span <7MB, increasing copy-number sensitivity for small regions across the genome could have great clinical value. The resolution limit of genome-wide copy-number variant (gwCNV) detection is driven by the sequencing depth and the distribution of FF in the patient population (e.g., it is more challenging to detect small deletions in samples with low FF). Attempting to increase resolution via deeper sequencing provides diminishing returns and quickly yields an economically inviable screening test. Therefore, methods to increase the FF, if feasible, are preferable.

Though FF may seem an immutable and intrinsic feature of a cfDNA sample, it can be altered, and strategies for increasing FF are revealed by factors that correlate with FF.^6,23,24^ For instance, FF is known to increase with gestational age,^7^ so drawing blood later in pregnancy leads to higher FF, though the effect is minor with FF increasing by <1% per week.^25,26^ FF also negatively correlates with first-trimester BMI and maternal age,^27^ but these values are effectively constant for any given pregnancy. At the molecular level, it has been observed that fetal-derived cfDNA fragments tend to be shorter,^28,29^ hypermethylated,^30-32^ and enriched at different locations than maternal cfDNA fragments.^33^ Leveraging these biases at the molecular and bioinformatic levels has the potential to multiplicatively boost the FF of every sample.

Here we present an analytical validation and extended characterization of a fetal-fraction amplification (“FFA”) technology that can be scalably applied to samples undergoing NIPS and yields significantly higher FF levels, thereby increasing sensitivity and specificity for all fetal anomalies arising from copy-number changes of any size across the genome.

## MATERIALS AND METHODS

### Ethics statement

All samples were from patients who had consented to de-identified research and received testing with the Prequel NIPS (Myriad Women’s Health, South San Francisco, CA; as described^9,34,35^). The study was granted an IRB exemption by Advarra (Pro00042194).

### FFA validation

The analytical validation of FFA involved 1,264 patient samples and 66 controls tested on 11 batches. Each patient sample was processed through two workflows: (1) standard WGS-based NIPS (i.e., Prequel without FFA) and (2) Prequel with FFA. The workflows were executed completely independently, each beginning with the extraction of cfDNA from replicate plasma aliquots. FF herein is measured either via a regression on autosomal bin depth^36^ or from the normalized depth of next-generation sequencing (NGS) data for a particular region (e.g., chrY, chr21, etc.). The proprietary FFA technology leverages the reduced size of fetal-derived cfDNA molecules—observed in several reports^28,29,37^—to increase the relative abundance of fetal cfDNA. Extracted cfDNA is quantified and then size selected by agarose gel electrophoresis such that the average length of selected cfDNA fragments is 140nt, which preferentially retains fetal cfDNA and depletes maternal cfDNA (Figure S1). The resulting libraries undergoing WGS have higher FF because fetal-derived fragments comprise a higher fraction of the total size-selected cfDNA.

Positive samples were sourced from our historical repository, prioritizing samples with confirmed clinical outcomes. The majority (81% of common aneuploidies and 55% overall) had orthogonally confirmed outcomes via diagnostic prenatal testing (e.g., amniocentesis or chorionic villus sampling) or diagnosis at birth. The minority without confirmed outcome were primarily screen-positive for microdeletions and RAAs on Prequel without FFA; thus, for these samples we are assessing comparability of Prequel with FFA to an already validated platform (i.e., Prequel without FFA). Negative samples were chosen randomly from the large population of patients who did not screen positive for any region of interest.

There were two types of negative-control samples used in validation and in every batch of samples screened in our clinical laboratory. First, in order to assess whether any contaminants were corrupting the steps in our DNA amplification, library preparation, and sequencing batch creation workflows, we included “no-template” controls (NTC) in which all steps were carried out as normal with the exception that no DNA was added to the DNA amplification. Next, to ensure that a euploid cfDNA sample indeed is identified as euploid by our pipeline, we included “pooled” controls. Pooled controls were either “XX” or “XY” and were created by pooling many hundreds of screen-negative samples with female or male fetuses, respectively.

Every batch of samples included in the validation contained NTCs and pooled controls, a plurality of negative samples, positives for each of the three common aneuploidies, and positives for some number of microdeletions, RAAs, and sex-chromosome aneuploidies (SCAs). Some individual samples were populated more than once in a single batch or in different batches to assess the intra- and inter-batch reproducibility, respectively.

To provide a more thorough characterization of FFA, some analyses herein augment the validation cohort with other samples tested internally during FFA development and verification. Only samples processed with the final, validated FFA protocol are included in such analyses.

### Sensitivity and specificity assessment

Because positive samples in analytical validation studies are relatively few in number and possibly unrepresentative (e.g., skewed toward above-average FF), using only the samples in the cohort to calculate sensitivity and specificity may not yield reflective estimates of clinical performance. Therefore, we developed a two-phase quantitative model that analyzes the positive samples included in the study but, importantly, overcomes the limitations of their rarity and potential biases. In the training phase of the model, a Markov-chain Monte Carlo analysis deciphers how z-scores of samples aneuploid for a given region (e.g., chromosome 21) scale as a function of FF and read depth. In the simulation phase, the model generates z-score distributions for an arbitrarily large number of mock samples; importantly, the mock positives are now relatively unbiased in key features like FF and sufficiently numerous to power accurate and low-error estimates of sensitivity and specificity. By iterating over many different z-score thresholds (i.e., the z-score cutoff between a positive and negative screening result) and calculating the sensitivity and specificity among the mock positives and negatives, the model yields a receiver-operator characteristic (ROC) curve. The ROC model described above can be applied to a group of regions (e.g., microdeletions) or a particular region (e.g., 22q11.2).

The ROC model is a principled and clinically reflective method of calculating sensitivity and specificity to overcome biases and sample-size limitations of a dataset, but we also calculated these metrics with the standard approach (see Supplement). Fetal genotype calls with the FFA protocol were classified as true positive (TP), true negative (TN), false positive (FP), and false negative (FN) based on their concordance with confirmed outcome, where available. For each class of aneuploidy—common aneuploidies, RAAs, SCAs, and microdeletions—sensitivity and specificity were calculated via standard definitions: sensitivity = TP / (TP + FN) and specificity = TN / (TN + FP).

The ROC model was not developed to apply to aneuploidy calls that rely on multiple regions simultaneously, such as the SCAs which rely upon coupled information from chromosomes X and Y; therefore, sensitivity and specificity values for SCAs were only calculated with the standard equations.

### Sex-call accuracy measurement

To determine the impact of FFA on distinguishing male and female fetuses, we calculated and compared the expected accuracy of sex calling for the standard-NIPS and FFA protocols. Sex calling is based on the FF estimated from chrY (FF_chrY_), with female-fetus pregnancies having FF_chrY_ ∼ 0 and male fetuses having FF_chrY_ > 0. A normal distribution was fit to FF_chrY_ data from pregnancies called as having female fetuses, and a beta distribution was fit to FF_chrY_ data from pregnancies called as having male fetuses. For a given sex-calling threshold, *y*, sex miscalls in female-fetus pregnancies are estimated by the amount of the normal-distribution fit with values exceeding *y*. Similarly, sex miscalls in male-fetus pregnancies were calculated as the share of the beta-distribution fit with values less than *y*. On the assumption that males and females are equally likely, a value of *y* was selected to minimize the total number of sex miscalls for both standard NIPS and FFA, and the difference in total expected FF-attributable sex miscalls was compared across protocols.

## RESULTS

### FFA increases FF an average of 2.3-fold for each sample

To directly measure the impact of FFA, we tested 2,401 samples from our verification and validation studies with both the standard-NIPS and FFA protocols, focusing particularly on the number of samples with FF<4%, the threshold for low FF suggested by American College of Medical Genetics and Genomics (ACMG).^38^ As shown in **Figure 1** (top), 3.7% of samples tested with the standard-NIPS protocol had FF less than 4%, whereas zero samples had low FF with the FFA protocol. The minimum FF observed in the FFA cohort was 4.9%. As it has been observed that samples from patients with high BMI tend to have low FF and cause elevated test failures on several NIPS platforms, we partitioned samples by their BMI classes (**Figure 1**, bottom). Critically, even at the highest BMI level (class III obesity), where 16% of samples had low FF with standard NIPS, every sample tested with the FFA protocol had FF > 4% (minimum FF observed among class III patients was 7.1%).

**Figure 1:**
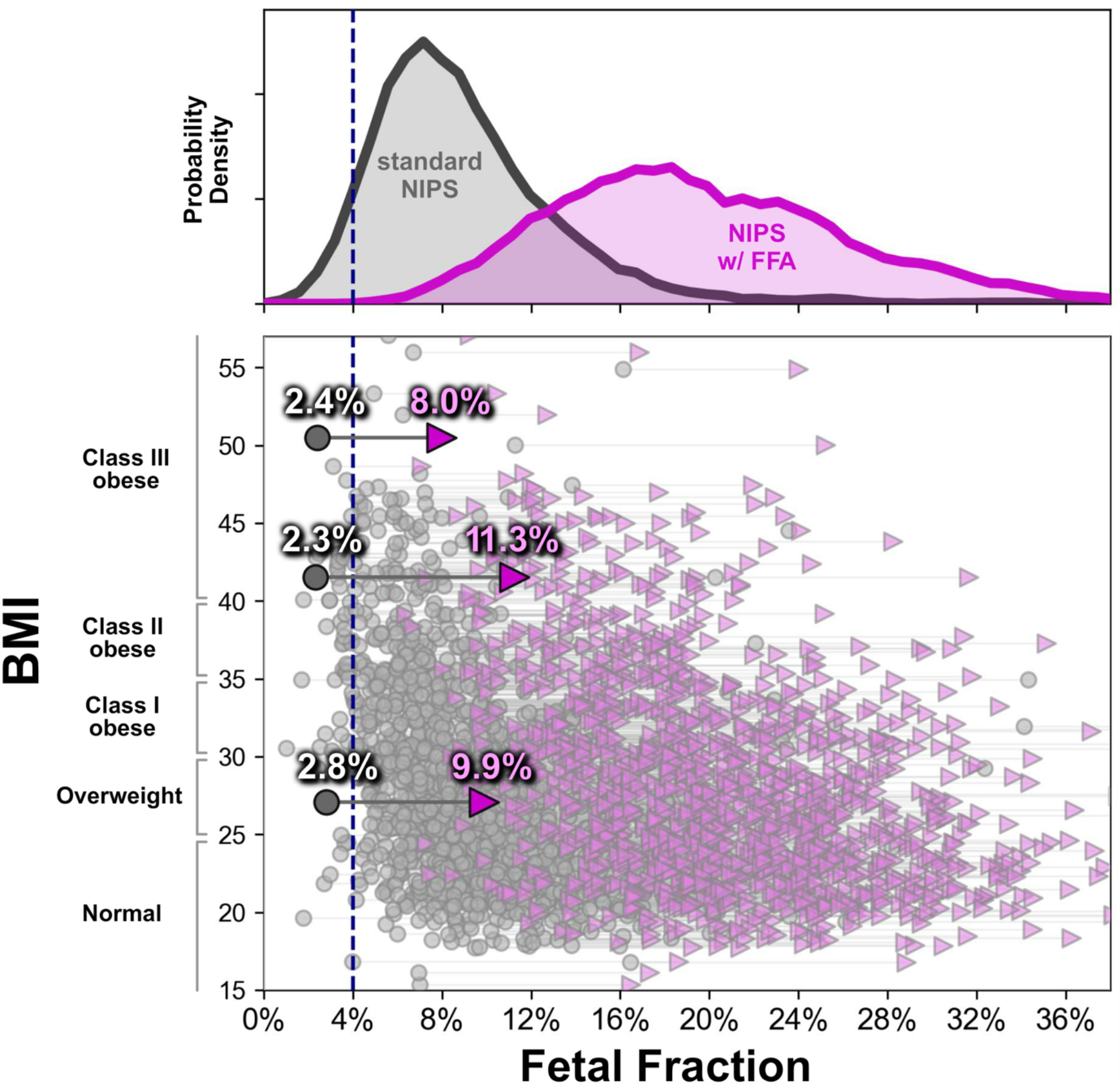
Fetal fraction amplification (FFA) technology increases fetal fraction (FF) across all BMI levels. For 2,401 patients who indicated BMI values on the test-requisition form, the FF levels measured before (gray circles) and after (purple triangles) FFA are plotted as a function of patients’ BMI values (vertical axis). The top panel plots the histogram of samples with or without FFA. The dotted line is at 4% FF, the threshold below which ACMG considers a sample to have low FF. Three samples are highlighted as illustrative examples that had low FF before FFA but normal FF after FFA.

To confirm that FFA did not artifactually increase FF by corrupting our FF-inference regression model (see Methods), we verified that the density of reads from chrY in pregnancies with male fetuses rose commensurately (**Figure S2**). We conclude that FFA increases FF by directly increasing the relative abundance of fetal-derived cfDNA fragments in each sequenced sample.

We examined sample-level changes in FF resulting from the FFA protocol because the upward shift in the overall FF distribution may obscure downward-shifting FF in a subset of samples. **Figure 2** shows the relative gain in FF conferred by FFA. Notably, 2,395 of the 2,401 samples tested (99.8%) had an increase in FF with FFA, with an average FF increase of 2.3-fold. The relative sample-level gain in FF varied as a function of FF (**Figure 2**): samples that were at low FF (<4%) with standard NIPS had the largest FF gain, with an average of 3.9-fold higher FF after undergoing FFA. Consistent with the FF gain diminishing at higher original FF levels, the six samples in which FF decreased with FFA had a median FF value of 27.8% (minimum 6.5%), and the FF with FFA remained high (median: 25.4%, minimum: 6.4%).

**Figure 2:**
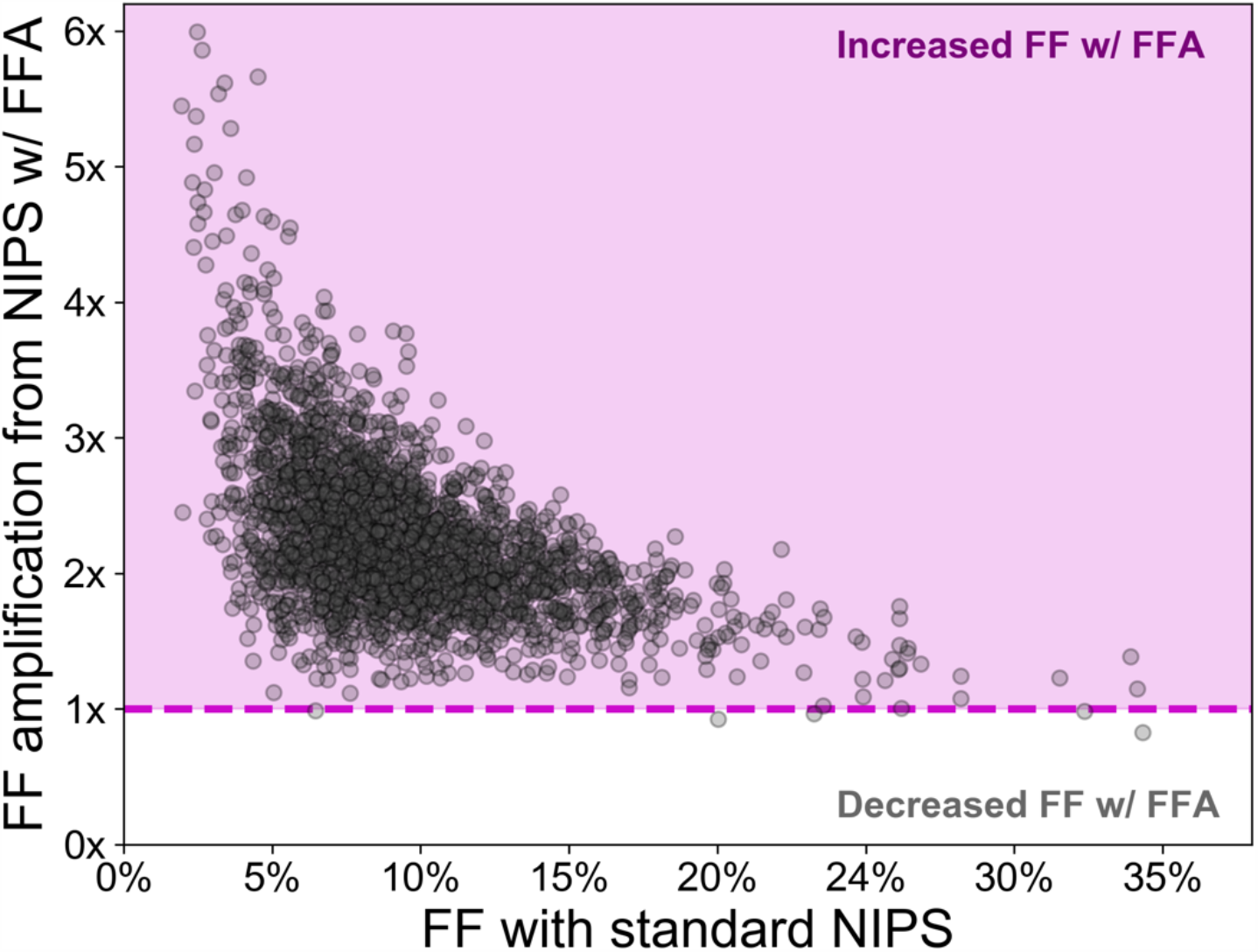
FFA increases FF for 99.8% of samples tested and most appreciably for low-FF samples. The fold-change difference in FF as a result of applying FFA is plotted for individual samples as a function of the original FF without FFA. The dashed line indicates no change in FF, and samples in the purple-shaded region had increased FF with FFA.

### FFA increases NIPS sensitivity for all regions of interest

In the same manner that FF can be directly measured in male-fetus pregnancies from the relative NGS depth of chrX and chrY, it is possible to measure FF of aneuploid samples via the relative NGS depth on the aneuploid chromosome (FF_positive_; **Figure 3A**). FF_positive_ is directly proportional to the z-score of an aneuploid region, and a higher z-score means that aneuploidy is more likely to be detected. Therefore, if FFA increases FF_positive_ of aneuploid regions, then FFA also increases NIPS sensitivity.

**Figure 3:**
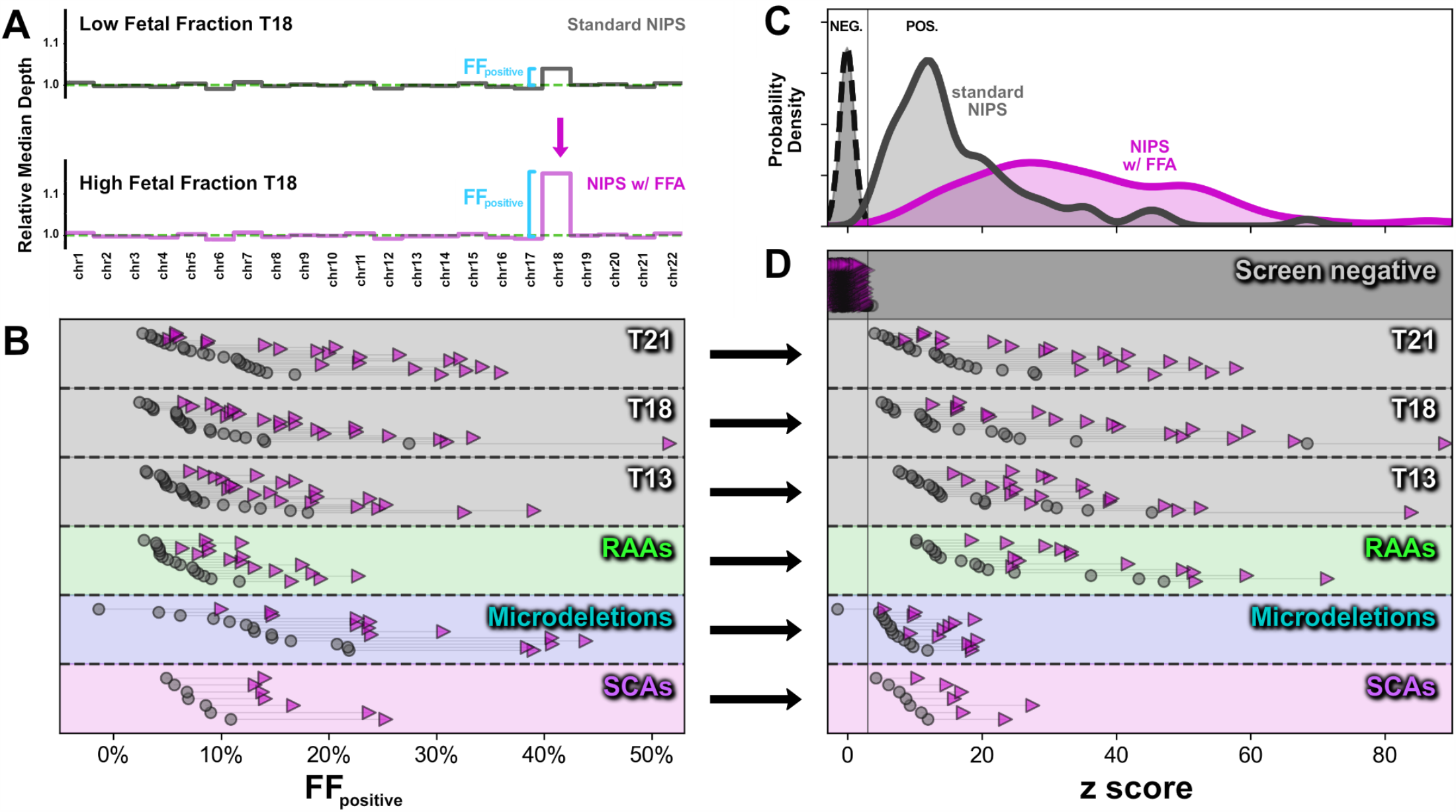
FFA improves detection of fetal chromosome abnormalities by amplifying the signal of aneuploid regions while maintaining background noise. **(A)** Schematic of the change in median depth per autosome as a result of FFA. The extent of the deviation from background is itself a measure of FF and is indicated as FF_positive_. **(B)** The increase in FF_positive_ without FFA (gray circles) and with FFA (purple triangles) is shown for aneuploid samples with the indicated chromosome anomalies. **(C and D)** z-scores without FFA (gray) and with FFA (purple) for the same samples as in (B) are stratified by their screening results and summarized either as population distributions (C) or as individual samples (D). For visual clarity in (C), the distribution of screen negative samples (“NEG.”; dashed line) has been scaled to be of comparable height as the screen-positive distributions to the right (solid lines). The vertical solid line indicates the z-score cutoff between screen-negative (left) and screen-positive (right) results. For SCAs, only female-fetus pregnancies are shown (i.e., MX and TX) because a z-score is used to identify chrX aneuploidies, whereas a two-dimensional analysis that does not use z-scores (not shown) is required for identification of XXY and XYY (FF_positive_ increased in all XXY and XYY pregnancies tested with FFA).

In every positive sample tested—across common aneuploidies, RAAs, and microdeletions—FFA yielded an increase in FF_positive_ (Figure 3B). As expected, FFA also increased z-scores for every tested aneuploid sample, whereas the z-score distribution for euploid samples was unchanged (**Figure 3C**,**D**). Larger z-score separation between positive and negative samples heightens the ability to discriminate such samples and thereby lessens the chances of false negatives and false positives. Together, these observations demonstrate that FFA directly increases the concentration of fetal-derived reads in each sample and enhances the sensitivity and specificity of NIPS.

A sample that screened negative for the 5p microdeletion with standard NIPS but positive with FFA (**Figure S3, Figure 3B**,**D**) provided further support for the enhanced sensitivity for fetal chromosome abnormalities that FFA confers. For this 3MB microdeletion, the copy number change was conspicuously apparent in the FFA data (**Figure S3**), converting a z-score below the calling threshold into one above the threshold (**Figure 3D**, microdeletions track).

To quantify the gain in sensitivity and specificity achievable with FFA, we analyzed the relationship between various clinical and technical metrics, such as z-scores, depth, incidence, and FF (see Methods). The ROC curves (**Figure S4**) for different classes of chromosomal abnormalities show that FFA enables near-perfect analytical sensitivity with near-perfect analytical specificity (**Table 1**). The sensitivity of common aneuploidies—shown to be high in our clinical experience without FFA^9^ — is marginally higher with FFA, as is the aggregate sensitivity of RAAs (**Figure S4**). However, the gain in microdeletion sensitivity is substantial: with FFA, the aggregate sensitivity for five common microdeletions is 97.2% at a joint specificity of 99.8%. For DiGeorge Syndrome in particular, FFA has an expected analytical sensitivity of 95.6% with an analytical specificity of 99.5%.

**Table 1:** Analytical performance metrics as estimated from ROC analysis.

|  | Analytical Sensitivity | Analytical Specificity |
| --- | --- | --- |
| Common aneuploidies (aggregate) | 99.988% ± 0.004% | 99.968% ± 0.005% |
| T21 | 99.990% ± 0.005% | 99.996% ± 0.001% |
| T18 | 99.990% ± 0.002% | 99.996% ± 0.001% |
| T13 | 99.978% ± 0.005% | 99.976% ± 0.005% |
| RAAs (aggregate) | 99.695% ± 0.305% | 99.981% ± 0.010% |
| Microdeletions (aggregate) | 97.172% ± 0.054% | 99.767% ± 0.012% |
| DiGeorge Syndrome (22q11.2) | 95.633% ± 0.071% | 99.949% ± 0.005% |

In addition to assessing performance with the ROC analysis above, we also observed that all samples with a confirmed aneuploidy or microdeletion were correctly identified with FFA **(Table S1**). Finally, the results were repeatable and reproducible within and across batches, respectively **(Table S2, S3**). Together, these experiments establish the analytical validity of FFA.

### FFA increases sex-calling accuracy relative to standard NIPS

Sex miscalls in NIPS arise from limitations that are either biological (e.g., true fetal mosaicism, vanishing twin) or technical (e.g., low FF). The former are an unavoidable aspect of NIPS on any screening platform (many sex miscalls occur at FF>>4%), but the latter could be mitigated by FFA due to its ability to increase the FF of all samples and thereby remove borderline calls. **Figure 4** shows distributions of FF_chrY_ (i.e., the FF as measured from the NGS read density on chromosome Y) for male-fetus and female-fetus pregnancies as observed for standard NIPS and FFA. Notably, the separation between male and female FF_chrY_ distributions is larger with FFA, reducing the chance of sex miscalls due to borderline FF_chrY_ values by an estimated 318x (see Methods). Underscoring the improvement, one sample tested in the validation study (**Figure 4**, orange arrow) was borderline in standard NIPS and miscalled as XX; however, the sample was clearly XY upon screening with FFA, and the pregnancy was orthogonally confirmed via ultrasound to be male.

**Figure 4:**
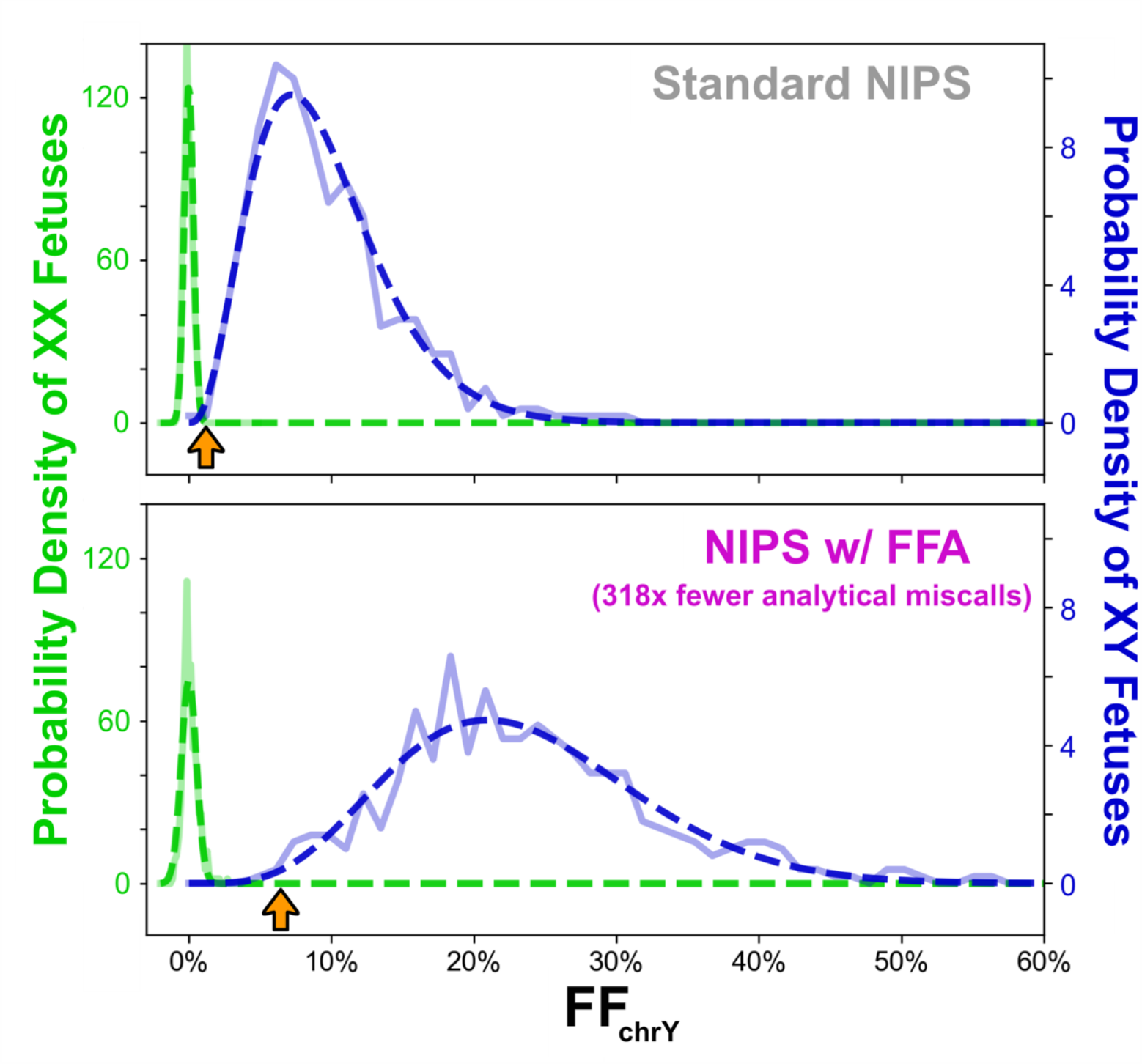
FFA increases the analytical accuracy of fetal sex calling. For both standard NIPS (top) and Prequel with FFA (bottom), the distribution of FF_chrY_ values is shown for samples called as female (green) or male (blue). Solid lines indicate raw data, and the dashed lines show best-fit traces for the female (Gaussian) and male (beta) populations. Only euploid samples are included in the analysis. The orange arrow depicts one sample tested on both platforms, called female in standard NIPS and male with FFA (the fetus was confirmed to be male). After minimizing the number of estimated miscalls on each platform (see Methods), analytical miscalls are predicted to drop 318-fold with FFA.

## DISCUSSION

Here we validated and characterized the performance of an NIPS that applies FFA technology to every sample. For 99.8% of samples tested, FF increased with FFA, with the average gain being 2.3-fold. Low-FF samples received the largest FF scaling, and out of 2,401 samples tested, 3.7% had low FF before FFA, but none had low FF after FFA. Importantly, the gain in FF is molecular and not algorithmic: FFA distinguishes between maternal and fetal DNA, and it increases the relative proportion of fetal DNA in the sample undergoing WGS. Though the combination of our custom algorithm and WGS technology showed high sensitivity and specificity for common aneuploidies across the FF spectrum without FFA technology,^9^ application of FFA increases performance for each type of aneuploidy, with the gain being particularly substantial for microdeletions.

The literature is replete with reports and professional-society statements expressing concern about low-FF samples.^8,38,39^ Many publications have explored and debated the merits of different approaches to handling low-FF samples: optimizing NIPS algorithms to issue confident results at low FF,^9^ failing such samples entirely,^40^ or pursuing mitigation strategies for failed low-FF samples, such as sequential redraw^41,42^ and fetal-fraction-based risk scores.^43,44^ However, consensus has remained elusive. As such, the FFA technology represents a paradigm shift in NIPS because samples that would have had low FF on standard NIPS are molecularly transformed into samples that have high FF. Accompanying the increase in test performance that FFA affords, we anticipate that this assay improvement will increase the confidence that providers and patients have in their results with NIPS.

FFA has a dramatic impact on the performance of microdeletion screening in NIPS. For common microdeletions, the expected aggregate sensitivity increases (Table 1, Figure S4), reaching 97.2% with FFA. In the past, when microdeletion sensitivity and specificity were low for microdeletions, ACOG recommended against microdeletion screening;^39^ however, we expect that sensitivity >97% and specificity >99% for microdeletions could allow professional societies to consider the clinical merits rather than the technological limitations of screening for microdeletions. Beyond the common microdeletions, our data suggest that FFA will increase the resolution of gwCNV detection, enabling confident identification of microdeletions below the current limit of 7MB achievable with standard NIPS. Notably, the 22q microdeletion, which causes DiGeorge Syndrome, most commonly spans ∼3MB and has an expected sensitivity of 95.6% with FFA. To ensure that false positives are rare, the resolution limit for novel gwCNV detection may need to be above 3MB, but dbVar^45^ contains more than a thousand unique pathogenic microdeletions between 3MB and 7MB in size, a number of which are associated with clinically serious phenotypes, so any gains in resolution should increase the utility of NIPS for patients and providers.

Even if two NIPS laboratories were to test the same plasma sample, the reported FF and sensitivity for aneuploidy may differ due to variations in the laboratories’ respective molecular and computational protocols. For instance, based on differing methods of aligning, filtering, counting, and analyzing NGS reads, a laboratory reporting 8% FF could have higher aneuploidy sensitivity than a laboratory reporting 10% FF. These differences complicate inter-lab comparisons of NIPS performance, especially since laboratories demonstrate performance on different sample sets and with different study designs (e.g., clinical experience study vs analytical validation study). As such, it can be difficult to make conclusive statements about relative NIPS performance. However, here we have demonstrated an unequivocal NIPS performance gain: two protocols (standard NIPS and FFA) were compared on a single set of samples within a single laboratory using a single aneuploidy-calling algorithm. FF increased 2.3-fold on average, and this FF increase resulted from a higher frequency of fetal-derived NGS reads. Beyond showing evidence for a relative gain in performance, the ROC analysis we performed yields an estimate of analytical sensitivity and specificity in an unbiased cohort reflective of a large population of clinical samples.

In conclusion, FFA fundamentally alters the landscape of NIPS by rendering a FF cutoff obsolete and negating the tradeoff often needed for a low test failure rate to coexist with highly accurate results. Debates about how best to serve patients with low-FF plasma samples can now be relegated to the past because low-FF samples need not exist.

## Data Availability

Certain data are available upon reasonable request to the corresponding author.

## Acknowledgements

The authors are grateful to Susan Hancock, Katherine Johansen Taber, and Anna Gardiner for assistance with the manuscript. Also, the authors acknowledge the support of the Myriad clinical laboratory in South San Francisco for processing the validation samples.

